# Longitudinal associations between physical activity and other health behaviours during the COVID-19 pandemic: A fixed effects analysis

**DOI:** 10.1101/2022.04.07.22273555

**Authors:** John J Mitchell, Feifei Bu, Daisy Fancourt, Andrew Steptoe, Jessica K Bone

## Abstract

**Background:** Government enforced restrictions on movement during the COVID-19 pandemic are likely to have had profound impacts on the daily behaviours of many individuals, including physical activity (PA). Given the pre-pandemic evidence for associations between PA and other health behaviours, changes in PA during the pandemic may have been detrimental for other health behaviours. This study aimed to evaluate whether changes in PA during and after the first national lockdown in the United Kingdom (UK) were associated with concurrent changes in other health behaviours, namely alcohol consumption, sleep, nutrition quality, diet quantity and sedentary time.

**Methods:** Data were derived from the UCL COVID-19 Social Study. The analytical sample consisted of 52,784 adults followed weekly across 22 weeks of the pandemic from 23rd March to 23rd August 2020. Data were analysed using fixed effects regression.

**Results:** There was significant within-individual variation in both PA and other health behaviours throughout the study period. Increased PA was positively associated with improved sleep and nutrition quality. However, increases in PA also showed modest associations with increased alcohol consumption and sedentary time.

**Conclusion:** Our findings indicate that, whilst the first wave of COVID-19 restrictions were in place, increases in PA were associated with improved sleep and better diet. Encouraging people to engage in PA may therefore lead to positive change in other health behaviours in times of adversity. However, increases in PA were also associated with more engagement in the negative health behaviours of alcohol consumption and sedentary time. These associations could be a result of increases in available leisure time for many people during COVID-19 restrictions and require further investigation to inform future public health guidance.

## Introduction

Physical activity (PA) supports numerous facets of lifelong health, including reducing morbidity and premature mortality [1, 2]. One way in which PA sustains good health may be through its associations with other health behaviours [3]. There is extensive evidence on the relationships between PA and other health behaviours, both from observational studies and from intervention studies testing PA as a way of supporting behaviour change. For instance, having a more physically active lifestyle has been linked to improved diet [3-6] and reduced alcohol consumption, by altering reward pathways [4, 7, 8] and promoting better self-regulation and decision-making [9, 10]. Increased PA is also related to better sleep quality [11-13] by altering circadian rhythms and mood as well as stress and thermoregulation pathways [14]. Moreover, there is also evidence that increases in PA are associated with reductions in sedentary time [2, 15, 16]. However, this relationship may not simply be inversely proportional as people may reallocate time from other daily PA into dedicated exercise PA instead of shortening their sedentary time [17-19]. Sedentary time also varies in forms, ranging from office work to television viewing, which may each show different relationships with PA [20]. Nonetheless, PA appears linked, to varying degrees, to other health behaviours through several, likely overlapping, physiological and psychosocial pathways [1, 3, 4, 21-26].

During the first year of the COVID-19 pandemic (2019/2020), lockdown measures were enacted by many countries to curb the spread of coronavirus by limiting peoples’ movements outside the home. During lockdowns, people were typically asked to shelter at home, with most workplaces, schools, and non-essential businesses (including gyms and playgrounds) closed for extended periods. Despite being effective in reducing virus transmission, lockdowns and other containment measures had a negative impact on certain health behaviours. Changes to PA were among the most acute of these negative impacts, with immediate changes to people’s daily lives following the introduction of ‘stay-at-home’ orders [27]. PA decreased during lockdowns globally [28-32], and there is some evidence of an inverse dose-response relationship between lockdown strictness and the number of steps people did per day [27].

Given the evidence for links between PA and other health behaviours, reductions in PA during the COVID-19 pandemic may have had negative impacts on other health behaviours. However, early evidence suggests that there has been extensive individual variation in changes in health behaviours during the pandemic, with studies reporting improvements in diet [33-40], alcohol consumption [41-43], and sleep for some individuals [35, 44, 45], despite evidence of population level decreases in PA, increases in sedentary time [27, 46, 47], alcohol consumption [41, 48, 49], and worse dietary choices [38]. The relationship between PA and these health behaviours is likely confounded by other major changes to people’s lives such as having to strictly isolate, family or financial adversity, children not being at school, or psychological distress in the wake of social restrictions [50]. People may have also used PA during lockdown to compensate for other unhealthy behaviours, as has been found in pre-pandemic studies exploring health behaviours [51, 52]. Although engagement in PA may have served as a protective factor against negative changes in other health behaviours, it is likely that these relationships differ compared to before the pandemic. Such change might be expected given the relationships between PA and other health behaviours not being previously explored at this extreme level of population movement restriction or in the context of the numerous other substantive changes to people’s lives during the pandemic. Few studies to date have explored within-individual changes in health behaviours during the COVID-19 pandemic, instead reporting population-level change [53-55] and, to our knowledge, no studies have yet explored changes in a range of health behaviours in the context of changes in individual PA levels. Understanding these links between health behaviours during the COVID-19 pandemic is of great clinical importance, both in terms of the longer-term health consequences of lockdowns and future periods of movement restriction. As new working patterns emerge in the wake of the COVID-19 pandemic, it is likely that behaviours associated with extended time at home may persist post-pandemic.

This study therefore aimed to explore these relationships during and after the first national lockdown in the UK. We used a large sample of 52,784 adults who were followed up weekly across 22 weeks between March and August 2020. Using fixed effects models, we investigated whether alcohol consumption, sedentary time, nutrition quality, diet quantity, and sleep showed parallel changes with changes in PA. We hypothesised that sedentary time would decrease, sleep quality would improve, nutrition quality would improve, and diet quantity and alcohol consumption would decrease concurrently with increases in daily PA. Testing these relationships will help us to understand the public health consequences of government-enforced lockdowns and the broader role PA may play in supporting positive changes in other health behaviours.

## Methods

### Participants

Data were drawn from the UCL Covid-19 Social study, a prospective panel study of more than 70,000 adults in the UK during the COVID-19 pandemic. Data were collected weekly online for a total of 22 weeks (between 21/03/2020 and 23/08/2020), then monthly thereafter. The study did not use a random sample design, but it does contain a heterogeneous sample that was recruited using three primary approaches. First, convenience sampling was used, including promoting the study through existing networks and mailing lists (including large databases of adults who had previously consented to be involved in health research across the UK), print and digital media coverage, and social media. Second, more targeted recruitment was undertaken focusing on (i) individuals from a low-income background, (ii) individuals with no or few educational qualifications, and (iii) individuals who were unemployed. Third, the study was promoted via partnerships with third sector organisations to vulnerable groups, including adults with pre-existing mental health conditions, older adults, carers, and people experiencing domestic violence or abuse. Ethical approval was provided by the UCL research ethics committee [12467/005]. All participants provided informed consent prior to enrolment. A full protocol for the study is available online at https://osf.io/jm8ra/.

Our study used weekly data during the first strict lockdown period in the UK (23/03/2020-10/05/2020) and the period of eased restrictions which followed until 23/08/2020. We included participants who provided a minimum of 2 repeated measures during the study period (N=60,132) as the fixed effects approach only models within-individual variation. We further excluded participants with missing data in any of the variables used in this study (12.2%). This yielded an analytical sample of 52,784 participants and 577,898 total observations (mean: 10.95 observations per individual) who contributed between 2 and 21 weeks of repeated measures.

### Measures

#### Physical activity

Participants were asked how many hours of the previous weekday were spent on i) *‘Going out for a walk or other gentle physical activity’*, ii) *‘Going out for moderate or high intensity activity (e*.*g*., *running, cycling or swimming)’*, and iii) *‘Exercising inside your home or garden (e*.*g*., *doing yoga, weights or indoor exercise)’*. Categories of response were ‘*did not do’, ‘<30 mins’, ‘30 mins-2 hours’, ‘3-5 hours’* or *‘*≥*6 hours’*. Mid-points of these categories were used to estimate participant’s total time participating in these three PA measures. Metabolic equivalence of task (MET) hours for each activity were calculated and then summed in line with the International Physical Activity Questionnaire scoring guidelines [56, 57] to create a final score of MET hours participating in dedicated PA. As such, an individual’s derived number of MET hours is a measure of not only the time they spent participating in activities, but also the metabolic requirements associated with those activities [58].

#### Health behaviours

Alcohol consumption was measured by asking participants, ‘*Over the past week have you drunk alcohol more than usual?’* with response options *‘less than usual’, ‘about the same’* or *‘more than usual’*, and a fourth option of abstinence. Responses were dichotomised into categories of *‘more consumption’*, or *‘less or the same consumption’*.

Sleep quality was measured by the question, ‘*Over the past week, how has your sleep been?*’. Participants selected from a 5-point scale with possible responses ‘*very poor’, ‘not good’, ‘average’, ‘good’ and ‘very good’*. Sleep quality was dichotomised into ‘*poor or average’* and ‘*good or very good’*.

Nutrition quality was measured with the question, *‘Over the past week how has your diet been?*’, with the responses being ‘*more healthy than usual’, ‘less healthy than usual’* or ‘*about the same healthiness as usual’*. This was also dichotomised as ‘*less healthy or the same’* or ‘*more healthy’*.

Diet quantity was reported in answer to the question, *‘Over the past week have you eaten more than usual?’*. Possible responses were eating *‘more than usual’, ‘less than usual’*, or *‘about the same as usual’*. Diet quantity was dichotomised as ‘*more or the same consumption*’ or ‘*less consumption’*, under the assumption that less consumption would represent a positive change in this variable.

Sedentary behaviour was measured as hours spent on various sedentary activities on the previous weekday. Participants were asked how long was spent watching television, browsing the internet, blogging, gaming, engaging in digital arts, reading, home-based creative tasks (such as painting, writing, playing music), and taking naps. Time participating in sedentary activities was measured using categorical responses: ‘*did not do’, ‘<30 mins’, ‘30 mins-2 hours’, ‘3-5 hours’* or *‘*≥*6 hours’*. Mid-points of each category were used to produce a continuous variable which indicated total hours spent participating in all sedentary activities.

#### Covariates

Covariates were also measured weekly and included time-varying measures of mental health and adversities. Depressive symptoms were measured with the Patient Health Questionnaire (PHQ-9) [59], with higher scores indicating more depressive symptoms, ranging from 0 to 27. Anxiety symptoms were measured using the Generalised Anxiety Disorder Assessment (GAD-7) [60], with higher scores indicating more symptoms of anxiety, ranging from 0 to 21. Adverse life events were also included, encompassing financial adversity such as being unable to pay rent or job loss, uncertainty in accessing medicines, and enforced self-isolation due to possible coronavirus infection, all of which were included as separate binary ‘yes’ or ‘no’ variables.

We also included baseline measures of time-invariant characteristics that may modify any identified relationships. These were gender (woman, man), age (under 50, ≥50 years old) and three self-reported health-related factors: i) any physical health condition (yes, no; including hypertension, diabetes, heart disease, lung cancer, or any other clinically diagnosed chronic physical health condition), ii) clinical diagnosis of a mental health problem (yes, no; depression, anxiety, or other) and iii) weight status (overweight [self-reported BMI ≥25] vs not overweight [BMI <25]).

### Statistical analysis

Data were analysed using fixed effects regression. This approach models only within-individual variation and thus automatically controls for observed and unobserved individual heterogeneity [61]. We first tested the longitudinal association between changes in MET hours of PA and changes in each health behaviour in separate fixed effects regression models. We then adjusted each model for time-varying confounders (depressive symptoms, anxiety symptoms, and adverse life events). Finally, we additionally adjusted each model for the other health behaviours. The final sample was weighted for analyses to the proportions of gender, age, ethnicity and education in the English population obtained from the Office for National Statistics [62]. Analyses were conducted using Stata 16 [63].

### Sensitivity analyses

First, we tested whether the association between PA and each health behaviour differed according to several demographic and clinical variables measured at baseline (gender, age, physical health conditions, clinically diagnosed mental health problems, and weight status). To do this, we included an interaction term between each of these variables and PA in separate fixed effects models. Second, to differentiate between the different components of sedentary time, we created two separate measures. One variable indicated time spent on screen-based sedentary activities (watching television, browsing the internet, blogging, or gaming) which were considered as sedentary behaviours bad for one’s health. Another variable indicated time spent on creative sedentary activities (engaging in digital arts, reading, home-based creative tasks), as sedentary time including engagement in the arts may still have some benefits for health [20, 64]. We then repeated analyses testing the association between PA and each of these types of sedentary time, before and after adjustment for the other type of sedentary time.

## Results

The analytical sample comprised 52,784 individuals, 75% of whom were women before weighting (Table 1). There was also an overrepresentation of white participants (95%) and participants with a degree or above (68%). After weighting, the sample reflected population proportions, with 50% women, 13% being ethnic minority, and 33% having a degree or above.

**Table 1.** Sample characteristics.

|  | Unweighted<br>N=52,784 | Weighted<br>N=52,784 |
| --- | --- | --- |
| **Age (mean, SD)** | 51.1 (14.7) | 46.7 (17.2) |
| **Women** | 76% | 52% |
| **White Ethnicity** | 95% | 86% |
| **Highest level of education** |  |  |
| No qualifications | 3% | 6% |
| Completed GCSE or equivalent (at school until age 16) | 11% | 26% |
| Completed post-16 vocational course | 6% | 9% |
| A-levels or equivalent (at school until age 18) | 12% | 26% |
| Undergraduate degree or professional qualification | 42% | 20% |
| Postgraduate degree | 26% | 13% |
| **What is your usual total household income per annum?** |  |  |
| <£16,000 | 15% | 21% |
| £16,000-£29,999 | 25% | 26% |
| £30,000-£59,999 | 35% | 33% |
| £60,000-89,999 | 15% | 12% |
| £90,000-119,999 | 6% | 4% |
| ≥£120,000 | 4% | 4% |
| **Time-varying sample characteristics at baseline** |  |  |
| **Physical activity, MET hours (mean, SD)** | 6.7 (9.3) | 6.8 (9.7) |
| **Alcohol consumption** |  |  |
| More than usual | 28% | 26% |
| **Sleep Quality** |  |  |
| Good or Very good | 34% | 32% |
| **Diet Quantity** |  |  |
| More than usual | 33% | 32% |
| **Nutrition Quality** |  |  |
| More healthy | 13% | 12% |
| **Sedentary time, hours (mean, SD)** | 7.0 (5.6) | 8.1 (6.0) |
| **Financial adversity** | 22% | 25% |
| **Difficulty accessing essentials** | 7% | 9% |
| **PHQ-9 Score (mean, SD)** | 7.2 (6.0) | 8.1 (6.7) |
| **GAD-7 Score (mean, SD)** | 5.45 (5.3) | 6.0 (5.8) |
| **Self-isolation due to COVID exposure** | 2% | 2% |
| **Time-varying Effect Modifiers at baseline** |  |  |
| **Clinically diagnosed mental health problem** | 20% | 22% |
| **Physical health condition** | 40% | 40% |
| **Overweight (BMI≥25)** | 55% | 58% |

On average across the study period, participants reported engaging in high levels of PA on the previous weekday (5.43 MET hours, standard deviation [SD]=8.18). This level of PA is equivalent to approximately 100 minutes of brisk walking, or 20 minutes of moderate to vigorous PA coupled with 60 minutes of walking. During the strict lockdown period (23/03/2020-10/05/2020), average reported PA was higher (5.77 MET hours, SD=8.44) than across the period of eased restrictions (10/05/2020-23/08/2020) that immediately followed (4.71 MET hours, SD=7.56). The most common type of PA in our sample was walking or other gentle physical activities (2.89 MET hours, SD=3.45), followed by exercising at home (1.32 MET hours, SD=2.75) and exercising outside (1.21 MET hours, SD=3.57). Meanwhile, participants reported doing sedentary activities for an average of 5.5 hours (SD=4.64) on the previous weekday. At baseline, 26% of participants reported consuming more alcohol than usual (Table 1). This varied over time, as within-individual variation in alcohol consumption accounted for approximately 43% of total variation. Sleep quality also showed substantial change, as within-individual variation accounted for 71% of the overall variation across the study period. However, within-individual variation accounted for relatively less of the overall variation observed in nutrition quality (31%) and diet quantity (34%) throughout the study period. Despite this within-individual variation, some participants did not demonstrate any change in the outcomes across the study period, meaning they were excluded from our fixed effects models. Nutrition quality was the most stable outcome, with the largest proportion of participants not demonstrating any change and were subsequently excluded from the fixed effects models (69%). Compared to the individuals included in the models, these participants were more likely to be male, older, have lower education, and report lower physical activity, but otherwise showed only small differences in outcomes and health-related factors (Supplementary Table 1).

### Fixed effects models

As shown in Table 2, we found that alcohol consumption increased in parallel with increased PA (coefficient [coef]=0.006, 95% confidence interval [CI]=0.003-0.010). This persisted after adjustment for time-varying confounders and other health behaviours (coef=0.007, 95% CI=0.003–0.009);. In contrast, sleep quality showed an improvement with increased PA (coef=0.016, 95% CI=0.012–0.019). This association was reduced, but not entirely attenuated, in the fully adjusted model (coef=0.010, 95% CI=0.007–0.014). There was no evidence that changes in diet quantity were associated with changes in PA. However, nutrition quality improved as PA increased, even after adjusting for other confounders and other health behaviours (coef=0.012, 95% CI=0.008-0.017). Lastly, increases in PA were associated with increases in time spent on sedentary activities, before and after adjustment for confounders and other health behaviours (coef=0.130, 95% CI=0.127-0.139).

**Table 2.** Fixed effects regression models testing within-individual time-varying associations between physical activity and health behaviours.

| Independent Variable: Metabolic Equivalent of Task Hours During Last Weekday |  |  |  |  |  |  |  |  |  |
| --- | --- | --- | --- | --- | --- | --- | --- | --- | --- |
| Health Behaviour | *Fixed Effects Model Controlling for time-invariant factors* |  |  | *Fixed Effects Model additionally adjusting for time-varying confounders* |  |  | *Fixed Effects Model additionally adjusting for time-varying confounders and other health behaviours* |  |  |
|  | Coef. | 95% C.I. | *p* | Coef. | 95% C.I. | *p* | Coef. | 95% C.I. | *p* |
| Alcohol Consumption | **0.006** | **0.003 - 0.010** | **<0.001** | **0.007** | **0.003 - 0.010** | **<0.001** | **0.006** | **0.003 - 0.009** | **<0.001** |
| Sleep Quality | **0.015** | **0.010 - 0.020** | **<0.001** | **0.014** | **0.010 - 0.017** | **<0.001** | **0.010** | **0.007 - 0.014** | **<0.001** |
| Diet Quantity | -0.002 | -0.007 - 0.002 | 0.314 | 0.000 | -0.005 - 0.039 | 0.850 | -0.003 | -0.008 - 0.002 | 0.227 |
| Nutrition Quality | 0.015 | **0.011 - 0.020** | **<0.001** | **0.013** | **0.010 - 0.018** | **<0.001** | **0.012** | **0.008 - 0.017** | **<0.001** |
| Sedentary Time | 0.133 | **0.130 - 0.143** | **<0.001** | **0.133** | **0.127 - 0.139** | **<0.001** | **0.130** | **0.127 - 0.139** | **<0.001** |

### Sensitivity analyses

Sensitivity analyses were conducted to assess whether the observed associations between PA and health behaviours differed according to a range of demographic and clinical variables. There was evidence that the relationship between PA and diet quantity varied by weight status (interaction term coef=0.012, 95% CI=0.001-0.024; Table 3). For those who were overweight, increased PA was associated with increased diet quantity (coef=-0.010, 95% CI=-0.019 - 0.001). In contrast, there was no evidence for any longitudinal association between PA and diet quantity in those who were not overweight (coef=0.002, 95% CI=-0.005 - 0.008). We found no evidence that any other factors (gender, age, clinically diagnosed mental health conditions, or physical health conditions) moderated associations between PA and other health behaviours.

**Table 3.** Interaction terms from fixed effects regression models testing whether the time-varying associations between physical activity and health behaviours differ according to demographic and health-related factors measured at baseline.

| Baseline Characteristic | Alcohol Consumption |  |  | Sleep Quality |  |  | Diet Quantity |  |  | Nutrition Quality |  |  | Sedentary Time |  |  |
| --- | --- | --- | --- | --- | --- | --- | --- | --- | --- | --- | --- | --- | --- | --- | --- |
|  | Coef | 95% C.I. | P-value | Coef | 95% C.I. | P-value | Coef | 95% C.I. | P-value | Coef | 95% C.I. | P-value | Coef | 95% C.I. | P-value |
| Gender*PA | -0.001 | -0.007 - 0.005 | 0.659 | -0.001 | -0.007 - 0.006 | 0.802 | 0.0003 | -0.009 - 0.01 | 0.956 | -0.001 | -0.007 - 0.005 | 0.802 | 0.011 | -0.001 - 0.217 | 0.063 |
| Age*PA | 0.003 | -0.003 - 0.010 | 0.313 | -0.002 | -0.009 - 0.005 | 0.498 | 0.0002 | -0.009 - 0.010 | 0.963 | -0.005 | -0.015 - 0.004 | 0.248 | -0.0002 | -0.013 - 0.012 | 0.976 |
| Clinically diagnosed mental health problem*PA | -0.003 | -0.012 - 0.005 | 0.450 | 0.003 | -0.008 - 0.013 | 0.648 | 0.005 | -0.005 - 0.016 | 0.757 | 0.0002 | -0.011 - 0.012 | 0.967 | 0.012 | -0.006 - 0.030 | 0.190 |
| Physical health conditions*PA | 0.002 | -0.005 - 0.008 | 0.641 | -0.004 | -0.011 - 0.003 | 0.292 | -0.004 | -0.014 - 0.005 | 0.387 | -0.002 | -0.011 - 0.008 | 0.693 | 0.0004 | -0.011 - 0.012 | 0.951 |
| Weight status*PA | 0.001 | -0.007 - 0.009 | 0.813 | -0.006 | -0.013 - 0.002 | 0.140 | 0.012 | 0.001 - 0.024 | 0.033 | 0.001 | -0.010 - 0.011 | 0.928 | 0.013 | -0.001 - 0.027 | 0.071 |

Finally, when examining the associations between PA and different types of sedentary time separately, increases in PA were associated with increases in both screen-based and creative sedentary activities (Supplementary Table 2).

## Discussion

In this study we tested the longitudinal associations of PA with other health behaviours, namely alcohol consumption, sleep quality, nutrition quality, diet quantity, and sedentary time during the COVID-19 pandemic. Increases in PA were associated with increases in alcohol consumption, sleep quality, nutrition quality, and time spent doing sedentary activities. However, changes in PA were not associated with changes in diet quantity. These associations were independent of all time-invariant confounders as well as a range of time-varying confounders and other health behaviours. There was limited evidence that the longitudinal association between PA and specific health behaviours differed according to baseline characteristics. Only participants who reported being overweight at the start of lockdown (according to their BMI) had significant increases in diet quantity in tandem with increases in PA, unlike participants who were not overweight.

Our sample showed a modest improvement in nutrition quality in parallel with increases in PA, although there were no associations between PA and diet quantity overall. Most studies have reported near-ubiquitous increases in snacking and overall food consumption during lockdowns [65-67]. This population-level increase in food consumption may have masked any possible associations between PA and diet quantity observed in this study. Nonetheless, the association of PA with improved nutrition is in line with pre-pandemic studies comparing the diets of active and inactive individuals, which report differences in food preferences; active individuals favour lower fat savoury foods despite having increased food demands overall [7, 8]. Mechanistically, acute exercise has been linked to reductions in levels of hormones controlling appetite (e.g., ghrelin) and may contribute to diet and nutrition changes, especially in response to stressful situations such as a lockdown [68, 69]. Worse food choices including snacking can also result from stress-induced cortisol spikes, and this spiking can be attenuated by acute bouts of PA [69]. Alongside existing evidence, our findings suggest that PA may have continued to help to support healthy dietary behaviours during the stresses of lockdown.

The relationship between increased PA and enhanced sleep quality observed in this study is likely bidirectional. Both single acute and regular bouts of exercise have been observed to improve individuals’ sleep by acting on a combination of circadian, metabolic, thermoregulatory, mood and endocrine pathways [11]. This is a positive feedback relationship, as good quality sleep supports both mood and daily energy levels which can indirectly improve engagement in PA [11]. It has previously been reported that altered work patterns during the COVID-19 pandemic led to widespread negative changes in sleep timings and increases in daytime napping for many, which may have longer-term negative consequences for psychological health [70]. Our findings suggest that encouraging PA could support sleep quality and counter this negative impact of the pandemic.

The relationship between increased PA and increased alcohol consumption in our sample may in part be attributable to the substantial overall increase in alcohol use in the population during the COVID-19 pandemic, coupled with changes in recreational time available [43, 49, 71, 72]. For many individuals, the increased leisure time available during the COVID-19 lockdown may have led to boredom, which may in turn have increased both PA and alcohol consumption. A recent study examining the motivations of adolescents to increase their PA during lockdown identified *‘increased time*’ and ‘*boredom*’ as substantial factors [73]. Boredom and the feeling of the slower passage of time have also been reported in at least one cohort during the pandemic [74]. Additionally, boredom has been associated with greater binge drinking, as well as both negative and positive changes to participant PA during the pandemic [48]. However, for others, increased free time may have been an opportunity and motivation to make positive lifestyle changes [75]. Differences in individuals’ backgrounds and circumstances likely influenced their health behaviours during lockdown. For instance, individuals with more available time due to working remotely or being on furlough may have chosen to spend their additional time in both positive and negative ways such as exercising, cooking more healthily, getting more sleep, but also consuming more alcohol and engaging in more sedentary activities, as seen in this study.

The observed association between increased PA and increased sedentary time is not surprising given the unique context of this study. In the enforced lockdown, sedentary hours may have replaced other low intensity PA such as walking to, from, and within work, taking children to school, or other outdoor chores and errands [17, 27, 31]. This increase in overall sedentary time is likely to have occurred for most people, even if they also did more dedicated PA (e.g., going on a walk or run, or working out at home) which is measured in this study. Additionally, increases in PA for exercise are often followed by increases in sedentary time in the following days due to acute fatigue and recovery [76]. This is most clear in exercise interventions for weight loss which often report a compensation phenomenon, either in participant’s diets or sedentary time, which partially offsets the benefits of increased PA [77]. This could be particularly relevant to individuals who used lockdown to improve their PA engagement. For example, participants in this study who reported being overweight had a greater increase in diet quantity with increased PA than participants who were not overweight. However, it is worth noting that the activities considered sedentary in this study are not intrinsically negative. Distinctions between positive and negative sedentary time have been drawn in previous literature which found differences in the associations of health outcomes with TV-viewing versus motorised commuting or sitting for work. Indeed, other evidence from this cohort has highlighted that some sedentary behaviours, such as remotely engaging in the creative arts, provided a way of coping with the stress of lockdowns [20, 64] and were associated with improvements in depression, anxiety and life satisfaction [78]. Our sensitivity analyses may support this finding, given that increases in PA were associated with increases in time spent on these creative arts activities. Nonetheless all types of sedentary time, even those that were screen-based and often considered negative, remained positively associated with PA.

This study has several strengths, including the use of a fixed effects approach and a large sample size with weekly measures across a period of 22 consecutive weeks during the first UK national lockdown and following easing of restrictions. However, it is important to acknowledge that findings from fixed effects models have limited generalisability to the population [61, 79], and the results of this study should be prefaced with *’within those whom change’*. Like other studies during the COVID-19 pandemic, there is a lack of pre-pandemic measures, limiting inferences regarding the magnitude of the associations observed in this study before versus during the pandemic. Further limitations include the use of a self-selected sample. Despite this, the large sample shows wide heterogeneity and good stratification across all major socio-demographic groups. Further efforts to improve sample representativeness included the use of weighting to align with national population statistics, making the sample comparable to the sample of another nationally representative study [80]. However, despite all efforts to make our sample inclusive and representative of the adult population, we cannot rule out the possibility of potential biases due to omitting other demographic factors that could be associated with survey participation in the weighting process. Additionally, our survey did not utilise a validated tool of daily PA, instead focussing on ‘dedicated’ or ‘leisure-time’ PA. Although this approach is widely used, it is less reflective of total daily movement. Lastly, we cannot rule out the possibility of reporting biases including social desirability in participant’s responses given the reliance on self-report tools, although use of a self-completed online survey may in-part have buffered this risk.

## Conclusion

This study of over 50,000 UK adults extends previous research on the changing PA behaviours of the UK population during the first national lockdown period [50] by asking whether these changes were linked to variations in other health behaviours. We identified a positive relationship between PA and improved nutrition quality and sleep quality throughout the first strict lockdown and period of eased restrictions. However, increased PA was also associated with increases in alcohol consumption and in sedentary behaviour. These findings may be explained by increases in available free time and changes to individual’s opportunities and motivations for behaviour change during lockdown. Future studies should explore which of these factors are most important, and the directionality of the observed relationships. Understanding the pathways by which these health behaviours are influencing one another, and the supporting role of PA, is vital for formulating guidelines for further lockdowns and the prevention of morbidity beyond the pandemic.

## Data Availability

Data are available upon reasonable request to the authors.

## Acknowledgements & Funding

This COVID-19 Social Study was funded by the Nuffield Foundation (WEL/FR-000022583), but the views expressed are those of the authors and not necessarily those of the foundation. The study was also supported by the MARCH Mental Health Network funded by UK Research and Innovation (ES/S002588/1), and by the Wellcome Trust (221400/Z/20/Z). DF was funded by the Wellcome Trust (205407/Z/16/Z). JJM is funded by an MRC grant (MR/N013867/1). The researchers are grateful for the support of several organisations with recruitment efforts, including the UKRI Mental Health Networks, Find Out Now, UCL BioResource, SEO Works, FieldworkHub, and Optimal Workshop. The study was also supported by HealthWise Wales, the Health and Care Research Wales initiative led by Cardiff University in collaboration with SAIL Databank developed by Swansea University.

