## Supplementary File for "Longitudinal associations between physical activity and other health behaviours during the COVID-19 pandemic: A fixed effects analysis"

### Outcome (MET hours) derivation:

Metabolic equivalence of tasks (MET) hours used based on Ainsworth et al., 2011 [1]. Purposeful leisure time physical activity measured, and their decided MET equivalent encompassed:

i) Exercising inside your home or garden (e.g. doing yoga, weights or indoor exercise) (4 METs; taken as the mid-point of the stated activities)
ii) Going out for moderate or high intensity activity (e.g running, cycling or swimming) (6 METs; taken as the mid-point of the stated activities)
iii) Going out for a walk or other gentle physical activity (3.3 METs)

Ainsworth, B.E., et al., *2011 Compendium of Physical Activities: a second update of codes and MET values.* Med Sci Sports Exerc, 2011. **43**(8): p. 1575-81.

Supplementary table 1. Comparison of sample characteristics at baseline of smallest sample included in fixed effects models (nutrition quality outcome models) due to invariance in the outcome.

| | Max excluded (Weighted) N=36,263 | Min Included (Weighted) N=16,521 |
|---|---|---|
| **Age (mean, SD)** | 47.1 (17.3) | 45.5 (16.7) |
| **Women** | 50% | 56% |
| **White Ethnicity** | 87% | 85% |
| **Highest level of education** | | |
| No qualifications | 7% | 4% |
| Completed GCSE or equivalent (at school until age 16) | 28% | 24% |
| Completed post-16 vocational course | 9% | 7% |
| A-levels or equivalent (at school until age 18) | 25% | 26% |
| Undergraduate degree or professional qualification | 19% | 22% |
| Postgraduate degree | 12% | 16% |
| **What is your usual total household income per annum?** | | |
| <£16,000 | 22% | 12% |
| £16,000-£29,999 | 26% | 22% |
| £30,000-£59,999 | 32% | 37% |
| £60,000-89,999 | 12% | 17% |
| £90,000-119,999 | 4% | 7% |
| ≥£120,000 | 4% | 5% |
| **Time-varying sample characteristics at baseline** | | |
| **Physical activity, MET hours (mean, SD)** | 6.6 (9.7) | 7.2 (9.6) |
| **Alcohol consumption** | | |
| More than usual | 27% | 25% |
| **Sleep Quality** | | |
| Good or Very good | 31% | 35% |
| **Diet Quantity** | | |
| More than usual | 32% | 35% |
| **Nutrition Quality** | | |
| More healthy | 3% | 3% |
| **Sedentary time, hours (mean, SD)** | 8.1 (6.0) | 7.9 (6.0) |
| **Financial adversity** | 25% | 26% |
| **Difficulty accessing essentials** | 9% | 7% |
| **PHQ-9 Score (mean, SD)** | 8.2 (6.7) | 8.0 (6.5) |
| **GAD-7 Score (mean, SD)** | 6.0 (5.9) | 6.0 (5.7) |
| **Self-isolation due to COVID exposure** | 2% | 2% |
| **Time-varying Effect Modifiers at baseline** | | |
| **Clinically diagnosed mental health problem** | 22% | 22% |
| **Physical health condition** | 41% | 37% |
| **Overweight (BMI≥25)** | 51% | 56% |

*ST1. Comparison of excluded and included participants in fixed effects models of nutrition quality due to invariance in nutrition quality during participants waves contributed during the study period.*

Supplementary table 2. Fixed effects regression models testing within-individual time-varying associations between physical activity, screen-based sedentary time, and creative sedentary time.

| | **Independent Variable: Metabolic Equivalent of Task Hours During Last Weekday** | | | | | |
|---|---|---|---|---|---|---|
| | ***Fixed Effects Model additionally adjusting for time-varying confounders, other health behaviours.*** | | | ***Fixed Effects Model additionally adjusting for time-varying confounders, other health behaviours and other sedentary activities.*** | | |
| **Health Behaviour** | **Coef.** | 95% C.I. | *p* | **Coef.** | 95% C.I. | *p* |
| Screen-based sedentary time | 0.093 | 0.089-0.098 | <0.001 | 0.076 | 0.072 - 0.080 | <0.001 |
| Creative sedentary time | 0.039 | 0.036-0.041 | <0.001 | 0.028 | 0.026 - 0.029 | 0.029 |

*ST2. Sensitivity fixed effects models of PA and sedentary activities when subdivided into those considered more 'positive' or 'enriching' than other screen-based sedentary activities.*
